# Impact of COVID-19 on the indigenous population of Brazil: A geo-epidemiological study

**DOI:** 10.1101/2021.01.12.21249703

**Authors:** Josilene D Alves, André S Abade, Wigis P Peres, Jonatas E Borges, Sandra M Santos, Alessandro R Scholze

**Affiliations:** Institute of Biological and Health Sciences. Federal University of Mato Grosso, Barra do Garças, MT, Brazil; Federal Institute of Education Science and Technology of Mato Grosso, Barra do Garças, MT, Brazil; Institute of Exact and Earth Sciences. Federal University of Mato Grosso, Barra do Garças, MT, Brazil; Faculty of Pharmaceutical. University Center of the Valley of Aragauaia, Barra do Garças, MT, Brazil; Faculty Nursing. State University of Northern Paraná, Paraná, Brazil

**Keywords:** indigenous, Brazil, SARS-CoV-2, vulnerability, spatial analysis

## Abstract

This study aimed to analyze the geographical distribution of COVID-19 and to identify highrisk areas for the occurrence of cases and deaths from the disease in the indigenous population of Brazil. This is an ecological study whose units of analysis were the Special Indigenous Sanitary Districts. Cases and deaths by COVID-19 notified by the Special Secretariat for Indigenous Health between March and October 2020 were included. To verify the spatial association, the Getis-Ord General G and Getis-Ord Gi ^*^ techniques were used. High spatial risk clusters have been identified by the scan statistics technique. 32,041 cases of COVID-19 and 471 deaths were reported. The incidence and mortality rates were between 758.14 and 18530.56 cases and 5.96 and 265.37 deaths per 100 thousand inhabitants, respectively. The non-randomness of cases (z-score = 5.40; p <0.001) and deaths (z-score = 3.83; p <0.001) was confirmed. Hotspots were evidenced for both events with confidence levels of 90, 95 and 99% concentrated in the North and Midwest regions of the country. Eight high-risk spatial clusters for cases with a relative risk (RR) between 1.08 and 4.11 (p <0.05) and two risk clusters for deaths with RR between 3.08 and 3.97 (p <0.05) were identified. The results indicate critical areas in the indigenous territories of Brazil and contribute to better targeting the control actions of COVID-19 in this population.

## INTRODUCTION

Coronavirus disease 2019 (COVID-19) is an infectious disease caused by severe acute respiratory syndrome coronavirus 2 (SARS-CoV-2) that was first identified in December 2019 in Wuhan, China, and it has spread globally causing a pandemic [1]. In Brazil surpassed 190000 people killed by Covid-19 in 2020, with a total of 7447625 people infected since the beginning of the pandemic. This fact places the country in the second place with the highest incidence of deaths caused by Convi-19, being surpassed only by the United States of America, which account for more than 330000 deaths [2].

According to Treweek et. al (2020) [3] the COVID-19 toll is not the same for everyone. Some studies show global evidence that elderly people, male, with comorbidities and in poverty are at a disproportionate risk of serious complications and deaths from COVID-19 [4,5]. Groups of ethnic minorities, who have these factors, have shown worse clinical results that include higher mortality and a higher rate of admission to Intensive Care Units [6]. 2020).

In Brazil, the Indigenous tribes have already seen their communities ravaged by exogenous diseases in the past. During the 2009 H1N1 influenza pandemic, their death rate was 4.5 times higher than the rest of the Brazil’s general population [7]. Even with the creation of the Emergency Plan to Combat COVID-19 in Indigenous Lands, through Law No. 14201, of June 7, 2020 [8], at the height of the pandemic that occurred in July 2020, mortality among indigenous peoples came to be 6.5 times greater than the rest of the population of Brazil [9,10].

In view of the failure to implement a reactive Emergency Plan that ignored the peculiarities already evidenced by other calamities that have decimated indigenous peoples throughout history, it is necessary to search for new strategies capable of assisting in the decision-making process to combat COVID-19 in indigenous lands.

Spatial analysis technologies have stood out in the context of the current pandemic for helping to understand the spatial and temporal dynamics of COVID-19, which is essential for their mitigation. Recognizing COVID-19 as a geographic phenomenon that is potentially mappable makes strategic planning and targeting of actions viable, as well as the formulation of more appropriate scientific and political responses[11,12].

Although geospatial tools are a strategic device, in terms of understanding situational diagnosis and monitoring disease progression, studies using these tools in the context of the indigenous population are still scarce [11,13].

Thus, considering the high degree of vulnerability of the indigenous population and the urgent need to identify priority territories, this study aimed to analyze the geographical distribution of COVID-19 and to identify areas of high risk for the occurrence of cases and death by disease in the indigenous population of Brazil.

## METHOD

### Study design and observation units

This is an ecological study using spatial analysis techniques[14], the units of which were the Special Indigenous Health Districts (SIHDs).

### Description of the study area and population

In Brazil, the self-declared indigenous population is estimated at 817000 individuals, representing 0.4% of the country’s total population. There are approximately 305 ethnic groups and 274 languages spoken, which represents one of the highest levels of sociodiversity in the world [15].

The indigenous health subsystem in Brazil is organized in 34 SIHDs and these areas are strategically divided by territorial criteria, based on the geographical occupation of indigenous communities. Its service structure has Indigenous Basic Health Units (IBHU), base centers and the Indigenous Health Support Houses (IHSH) [16]. Figure 1 presented the location of each SIHD.

**Figure 1:**
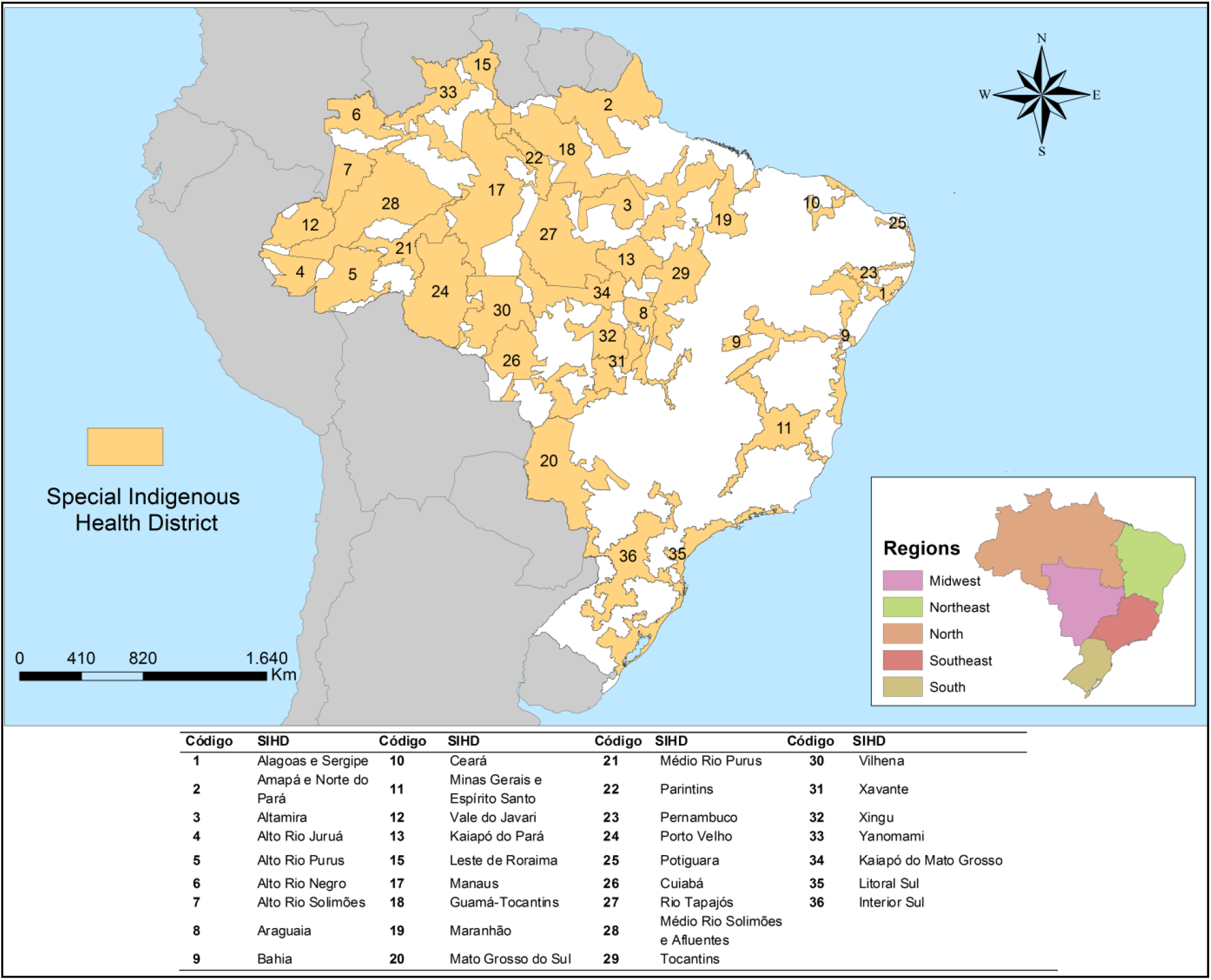
Map of Special Indigenous Health Districts and their geographical location. Brazil, 2020.

The study population consisted of cases with confirmed diagnosis and deaths by COVID-19 reported in the Brazilian indigenous population from March 24 to October 26, 2020. It is noteworthy that confirmed cases are characterized as cases with a positive result by laboratory confirmation or confirmed by clinical, clinical-epidemiological and clinical-image criteria [10].

### Study information sources

The study used the free access epidemiological bulletins published by the Special Secretariat for Indigenous Health (https://saudeindigena.saude.gov.br/corona), the institution responsible for coordinating and implementing the National Health Care Policy on Indigenous Peoples in the national territory. The data refer to the situation of the coronavirus in indigenous people served by the Indigenous Health Care Subsystem in each of the 34 SIHDs [10].

### Data Analysis

First, we conducted an exploratory analysis of the cases and deaths that occurred in the period, calculating the absolute number and the incidence and mortality rates according to each month investigated. Then, the same calculations were performed according to sex and age group. To calculate the incidence and mortality rates, the ratio between the total number of cases or deaths that occurred during the study period and the total indigenous population was used. The result was subsequently multiplied by 100000. The same rates were calculated by SIHD, using the number of cases or deaths recorded in each SIHD and the indigenous population of each SIHD.

### Spatial analysis

To verify the spatial association of the incidence and mortality rates by COVID-19, were used techniques Getis-Ord General G and Getis-Ord Gi^*^[17]. The Incremental Spatial Autocorrelation tool in the ArcGis V.10.5 software was used to obtain the best distance at which the spatial clusters were most pronounced [18].

The High/Low Clustering (Getis-Ord General G) statistic tool gives a general analysis of the area.The null hypothesis states that there is no spatial clustering of feature values. When the p-value returned by this tool is small and statistically significant, the null hypothesis can be rejected. Case the null hypothesis is rejected, the sign of the z-score becomes important, where their values of ± 3 represent a 99% confidence level. If the z-score value is positive, the observed General G index is larger than the expected General G index, indicating that high values for the attribute are clustered in the study area. If the z-score value is negative, the observed General G index is smaller than the expected index, indicating that lower values are clustered in the study área [19].

To examine spatial patterns in detail, the hot spot technique (Getis-Ord Gi^*^) was used, which analyzes the spatial association locally, based on a spatial concentration indicator. In Getis-Ord Gi^*^ the values for each locality, that is, each SIHD, are considered from a neighborhood matrix. In this technique, statistically significant spatial clusters of high values (Hotspot) and low values (Coldspot) are identified [17].

This analysis also generates a z-score for statistically significant census sectors. The higher the z-score, the more intense the grouping of high values (Hotspot), while the lower the value, the more intense is the grouping of low values or less occurrence of the event (Coldspot). In addition to the z-score, the p-value and significance level (Gi-Bin) are provided [17].

The technique of spatial scan statistics developed by Kulldorff and Nagarwalla (1995)[20] was used to identify areas at risk of COVID-19-related occurrences and deaths.

Scan statistics are performed through windows that move along the study territory around the centroid, corresponding to the centre of each analyzed territorial unit (SIHD). The formation of spatial clusters is based on the calculation of the number of events found within each window [21].

The likelihood function is maximized over all window locations and sizes, and the one with the maximum likelihood is the most likely cluster. The likelihood ratio (LLR) for this window is the test statistic for the maximum LLR. In addition to identifying spatial clusters, the method provides the relative risk (RR) which is a value that represents how much an area is more or less susceptible to having the presence of the event in relation to the other areas of the entire studied territorial extension [21].

Thus, the null hypothesis was that the number of cases and deaths observed are geographically distributed at random. And the alternative hypothesis was that there is some agglomeration, that is, the cases and deaths from COVID-19 observed are higher than expected. For the analysis of this study, the following criteria were adopted: identification of essentially high-risk spatial clusters, use of the discrete Poisson model with no geographical overlap of the clusters, circular shaped clusters and 999 Monte Carlo replications. The size of the exposed population stipulated by the Gini coefficient [22]. A p value <0.05 was adopted for statistically significant clusters and the 95% confidence interval (CI) was estimated.

### Software

The tabulation and descriptive analysis of the data set used spreadsheet software. SatScan® software version 9.6 was used for the spatial scan statistic technique. The spatial association and the construction of maps were performed using ArcGis® software version 10.6.

### Ethical considerations

Because it is research with secondary data of public access using data in an aggregated form and without the nominal identification of the subjects, the opinion by a Research Ethics Committee was waived, according to the National Health Council Resolution 510/2016.

## RESULTS

During the study period, 32041 cases of COVID-19 and 471 deaths were reported. Women represented 52.10% (n = 16686) of reported cases and 49.29% (n = 15792) of cases were in the 20-49 age group. Regarding deaths, 66.24% (n = 312) occurred among men and the most affected age groups were 50 to 79 years old (50.11%; n = 236) and aged over 80 years (34.18%; n = 161). When analyzing the incidence and mortality rates, we show that both cases and deaths increase with the age of the indigenous population regardless of gender (Figure 2).

**Figure 2:**
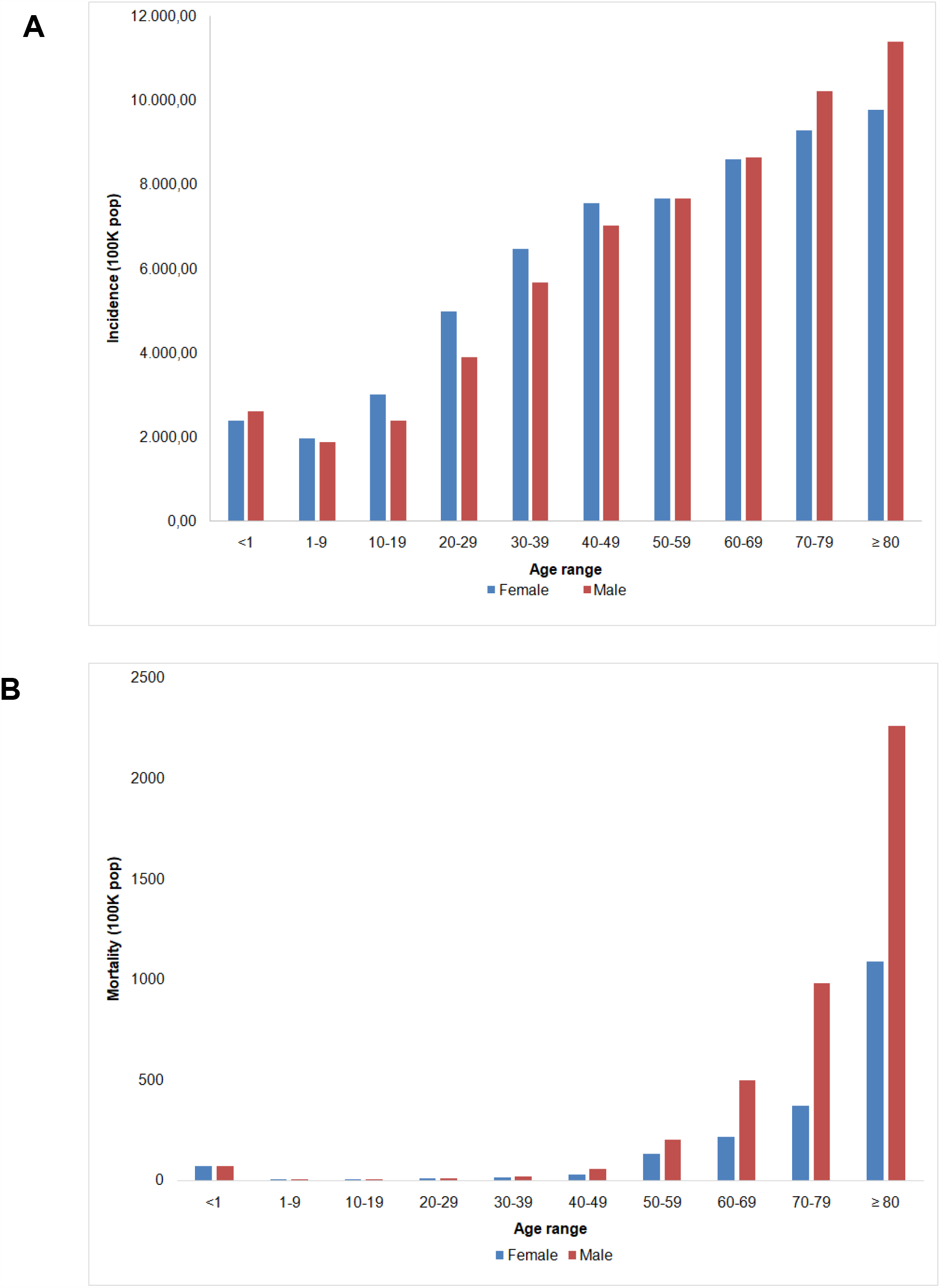
Distribution of the incidence rate (A) and mortality rate (B) by COVID-19 among the indigenous population according to sex and age group. Brazil, 2020.

In Figure 3 (A-B) we presented the temporal evolution of cases and deaths by COVID-19 among the indigenous population and their respective population rates per 100000 inhabitants. We observed that the number of cases and deaths has been increasing since the beginning of the pandemic in March 2020, with the peak occurring after five months, in July 2020.

**Figure 3:**
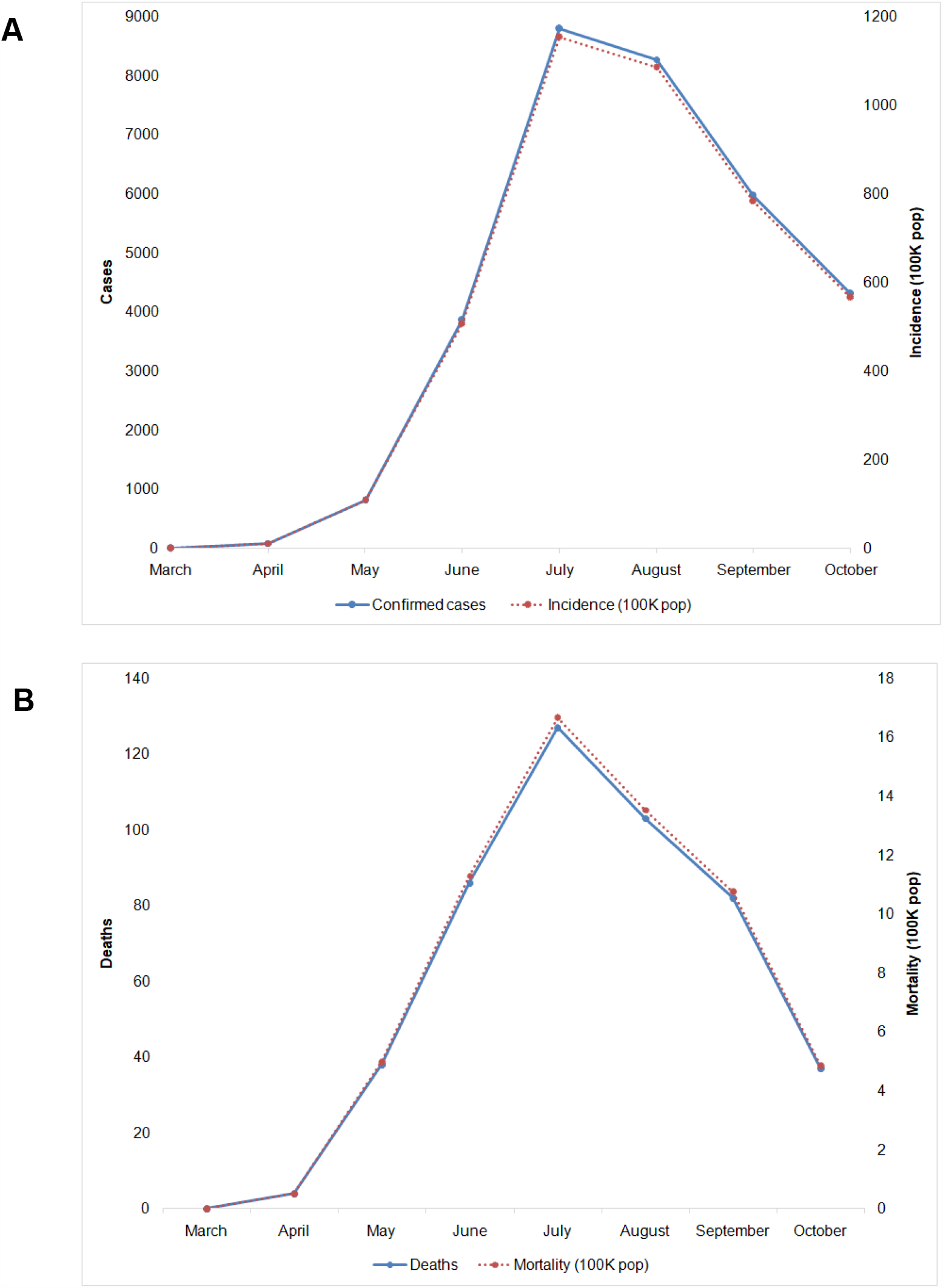
Temporal evolution of cases (A) and deaths (B) by COVID-19 among the indigenous population and their respective population rates per 100 thousand inhabitants. Brazil, 2020.

The distribution of incidence and mortality rates by COVID-19 for each SIHD are presented in Figure 4 A and B, respectively. When analyzing the incidence of the disease, it was found that the incidence rate varied between 758.14 and 18530.56 cases per 100000 inhabitants, being that the SIHDs located in the Midwest and North regions were the most affected. The SIHDs with the highest incidence were: Kaiapó do Pará, Altamira, Cuiabá, Kaiapó do Mato Grosso and Rio Tapajós. In relation to mortality, the rate varied between 5.96 and 265.37 deaths per 100000 inhabitants. The North, Midwest and South regions had the highest rates. The SIHDs with the highest mortality rates were: Cuiabá, Vilhena, Xavante, Xingu and Kaiapó do Pará.

**Figure 4:**
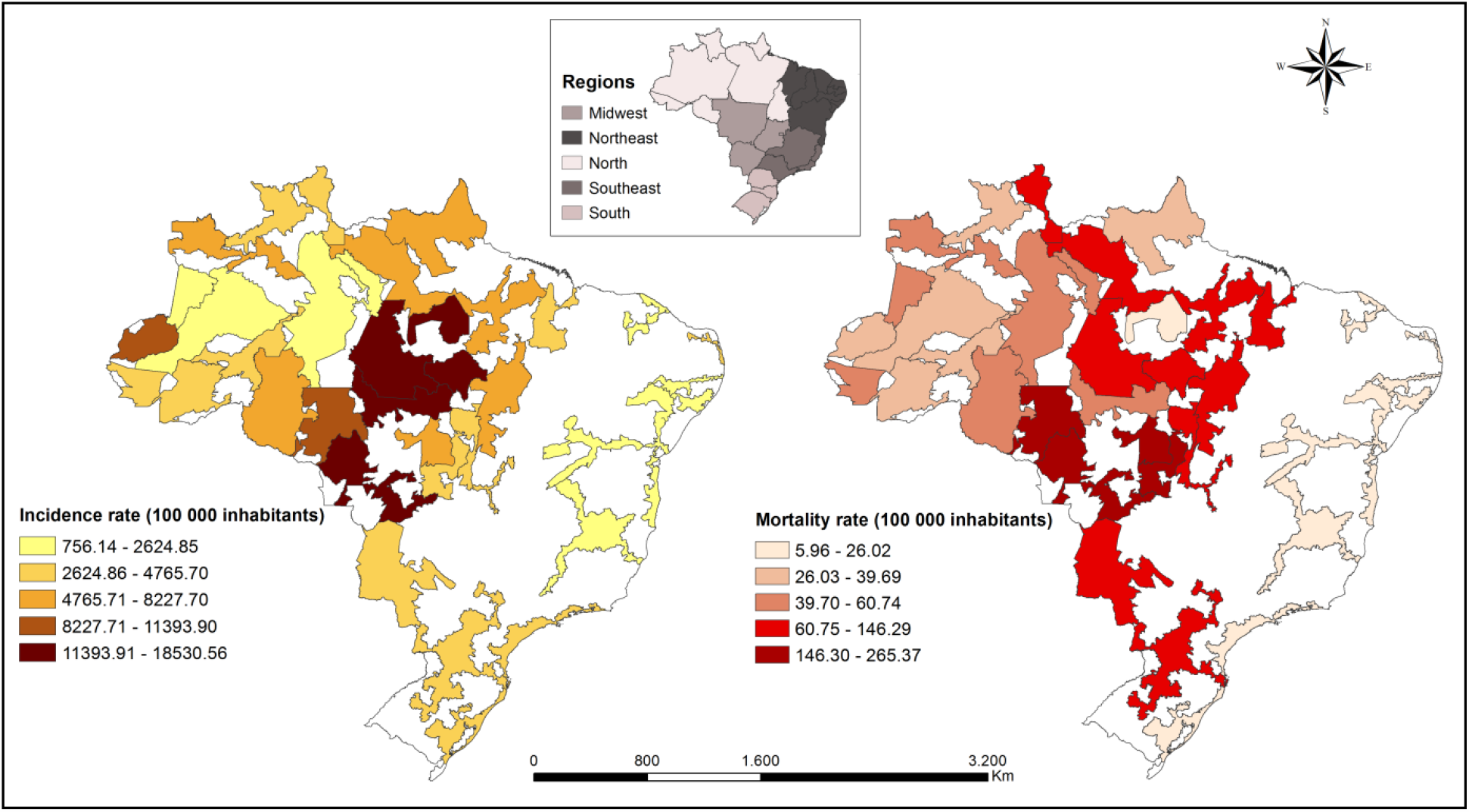
Distribution of incidence (A) and mortality (B) rates to COVID-19 in Special Indigenous Health District. Brazil, 2020.

The G Getis-Ord technique presented in Figure 5A, points to a z-score of 5.40 whose pseudo-significance test value was p <0.001, which shows the non-randomness of COVID-19 cases since the distribution high spatial values in the data set are spatially more grouped than expected indicating the formation of clusters. Figure 5B shows the local spatial association of the COVID-19 incidence rate, using the Getis-Ord Gi^*^ technique, which allowed the identification of hotspots formed by SIHD located in the Midwest and North regions of Brazil with confidence levels 90 and 99%. In the hotspots are the areas with the largest group of high values for the incidence of the disease.

**Figure 5:**
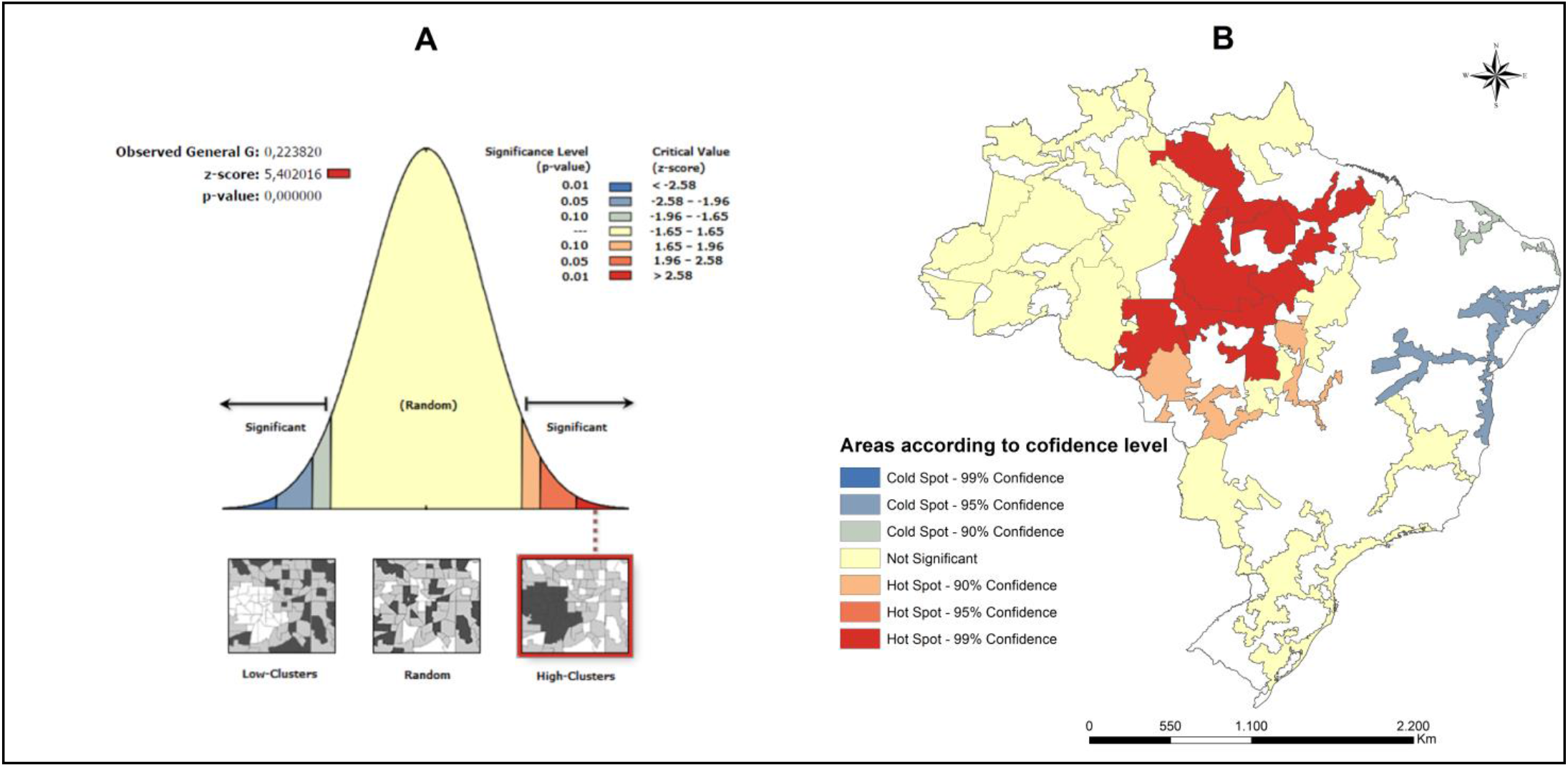
Hotspots and coldspots for COVID-19 cases in Special Indigenous Sanitary Districts. Brazil, 2020. (A) Level of statistical significance of Getis-Ord G for COVID-19 cases. (B) Spatial clusters of COVID-19 cases according to the level of confidence.

Figure 6A shows the result of the Getis-Ord General G technique (z-score = 3.83; p <0.001), which indicates the non-randomness of deaths by COVID-19 and the presence of spatial clusters. The Getis-Ord Gi^*^ technique showed hotspots with confidence levels of 90, 95 and 99% concentrated mainly in the Midwest region of the country.

**Figure 6:**
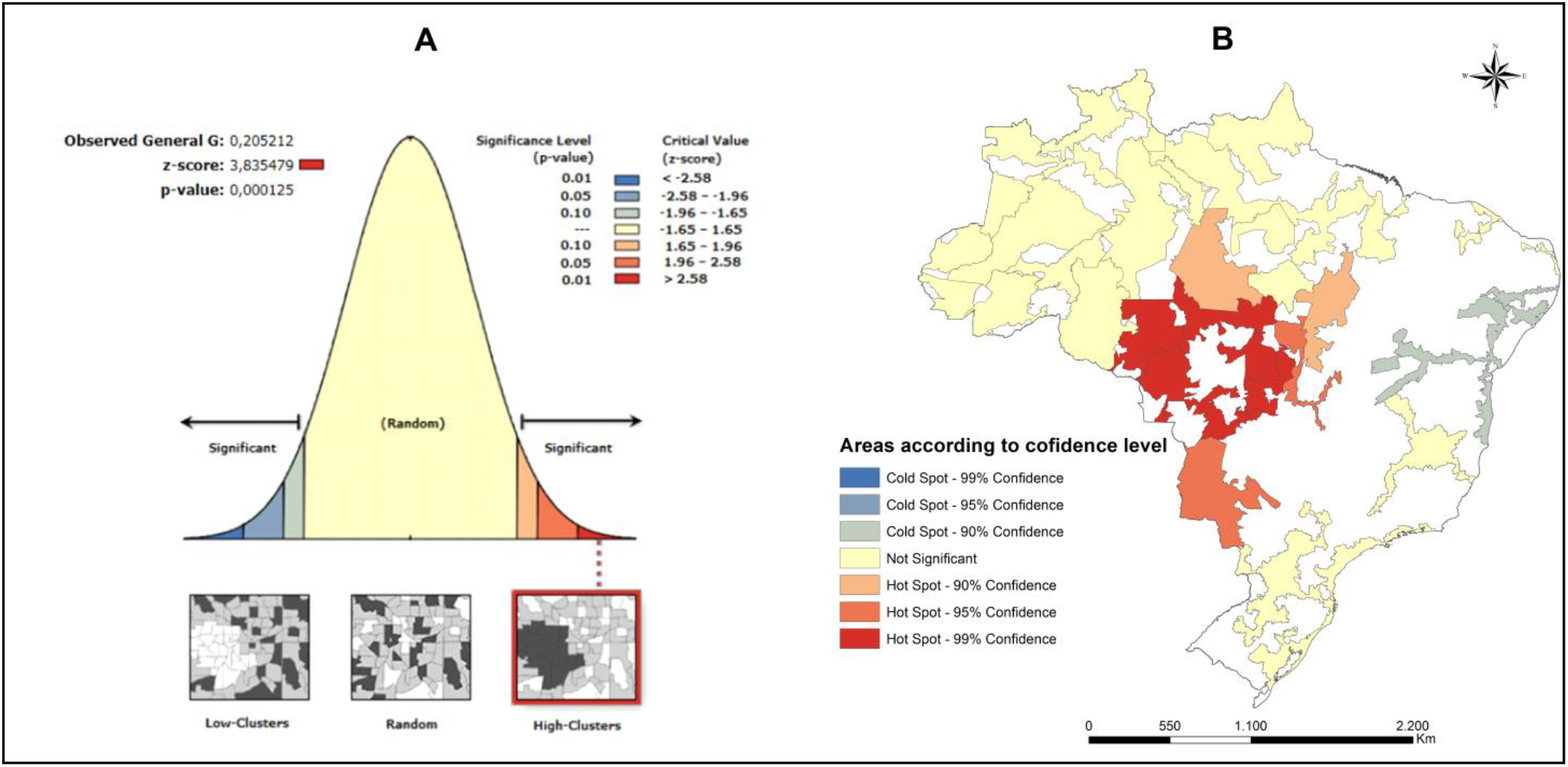
Hotspots and coldspots for COVID-19 deaths in Special Indigenous Sanitary Districts. Brazil, 2020. (A) Level of statistical significance of Getis-Ord G for COVID-19 cases. (B) Spatial clusters of COVID-19 deaths according to the confidence level.

In the northeastern region of the country, coldspots were identified, which are groups with low values, both for the occurrence of cases (Figure 5B) and deaths (Figure 6B) by COVID-19.

When applying the scan statistic, 8 high-risk spatial clusters were identified for the occurrence of COVID-19 cases that were statistically significant. In total 14 DSEI were part of the risk areas and were located in the north, central west, south and southeast regions (Figure 7). The largest spatial cluster of risk was formed by the DSEIS Altamira, Kaiapó of Pará, Rio Tapajós and Kaiapó of Mato Grosso. The population at risk in this cluster was 28,693 indigenous people and the RR was 4.11 (p <0.001), which indicates that indigenous people from these DSEI are 4 times more likely to become infected with COVID-19 compared to those who do not belong to these locations. The characteristics of the other risk spatial clusters identified are specified in Table 1.

**Table 1:** Characteristics of high-risk space clusters for the occurrence of cases of COVID-19 in the indigenous population.

| **Risk Cluster** | **Spatial units** | **Population at risk** | **LLR** | **Relative risk** | **p-value** | **95% IC** |
| --- | --- | --- | --- | --- | --- | --- |
| 1 | Altamira, Kaiapó do<br>Pará, Rio Tapajós<br>and Kaiapó do Mato<br>Grosso | 28693 | 2729.28 | 4.11 | <0,001 | 2.02-2.15 |
| 2 | Cuiabá and Vilhena | 14600 | 959.86 | 3.34 | <0,001 | 2.00-2.15 |
| 3 | Amapá and Norte do<br>Pará and Guamá-Tocantins | 30162 | 365.79 | 1.89 | <0,001 | 2.01-2.18 |
| 4 | Vale do Javari | 6281 | 223.33 | 2.58 | <0,001 | 1.94-2.25 |
| 5 | Alto Rio Negro | 28858 | 119.98 | 1.49 | <0,001 | 1.99-2.19 |
| 6 | Maranhão and<br>Tocantins | 49698 | 45.52 | 1.23 | <0,001 | 2.00-2.17 |
| 7 | Interior Sul | 38945 | 14.78 | 1.14 | <0,001 | 1.99-2.19 |
| 8 | Manaus | 51797 | 6.58 | 1.08 | 0.014 | 2.01-2.18 |

Source: Elaborated by the authors. LLR: Likelihood ratio; 95% CI: 95% confidence interval.

**Figure 7:**
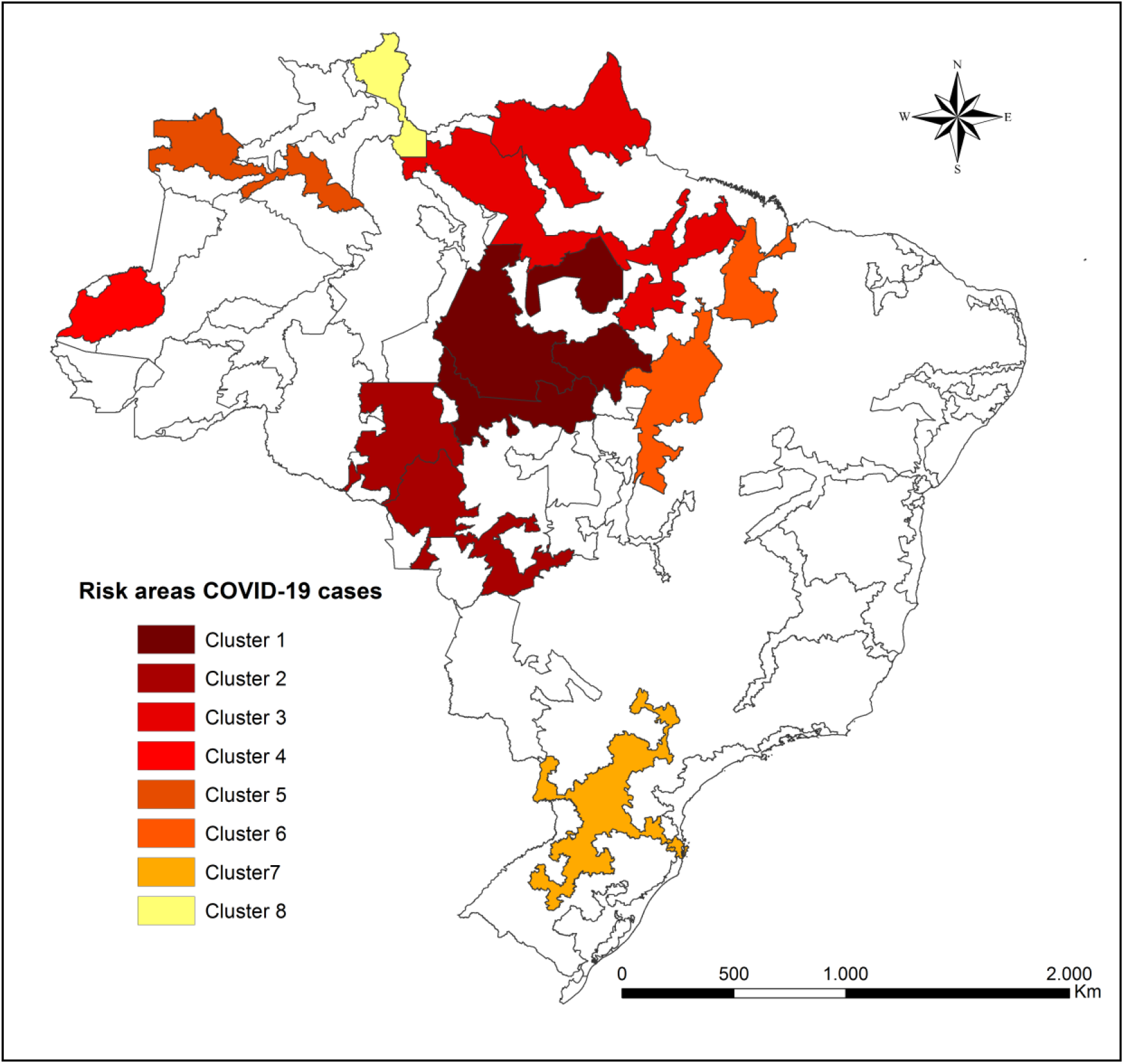
Spatial risk areas for the incidence of COVID-19 in the indigenous population according to a Special Indigenous Health District. Brazil, 2020.

In Figure 8 are presented the two high-risk spatial clusters identified for mortality from COVID-19 that are located in the central west region of the country. Cluster one includes the SIHDs Cuiabá, Vilhena and Xingu and presents RR = 3.97 (p <0.001). Cluster two is formed by the SIHDs Araguaia and Xavante and has RR = 3.08 (p <0.001). In total, the two clusters have a population of 50323 indigenous people at risk of mortality from COVID-19 (Table 2).

**Table 2:** Characteristics of high-risk space clusters for COVID-19 mortality in the indigenous population.

| Risk Cluster | Spatial units | Population at risk | LLR | Relative risk | p-value | 95% IC |
| --- | --- | --- | --- | --- | --- | --- |
| 1 | Cuiabá, Vilhena and Xingu | 22600 | 30.56 | 3.97 | <0.001 | 1.58-2.75 |
| 2 | Araguaia and Xavante | 27723 | 20.76 | 3.08 | <0.001 | 1.57-2.77 |

Source: Elaborated by the authors. LLR: Likelihood ratio; 95% CI: 95% confidence interval.

**Figure 8:**
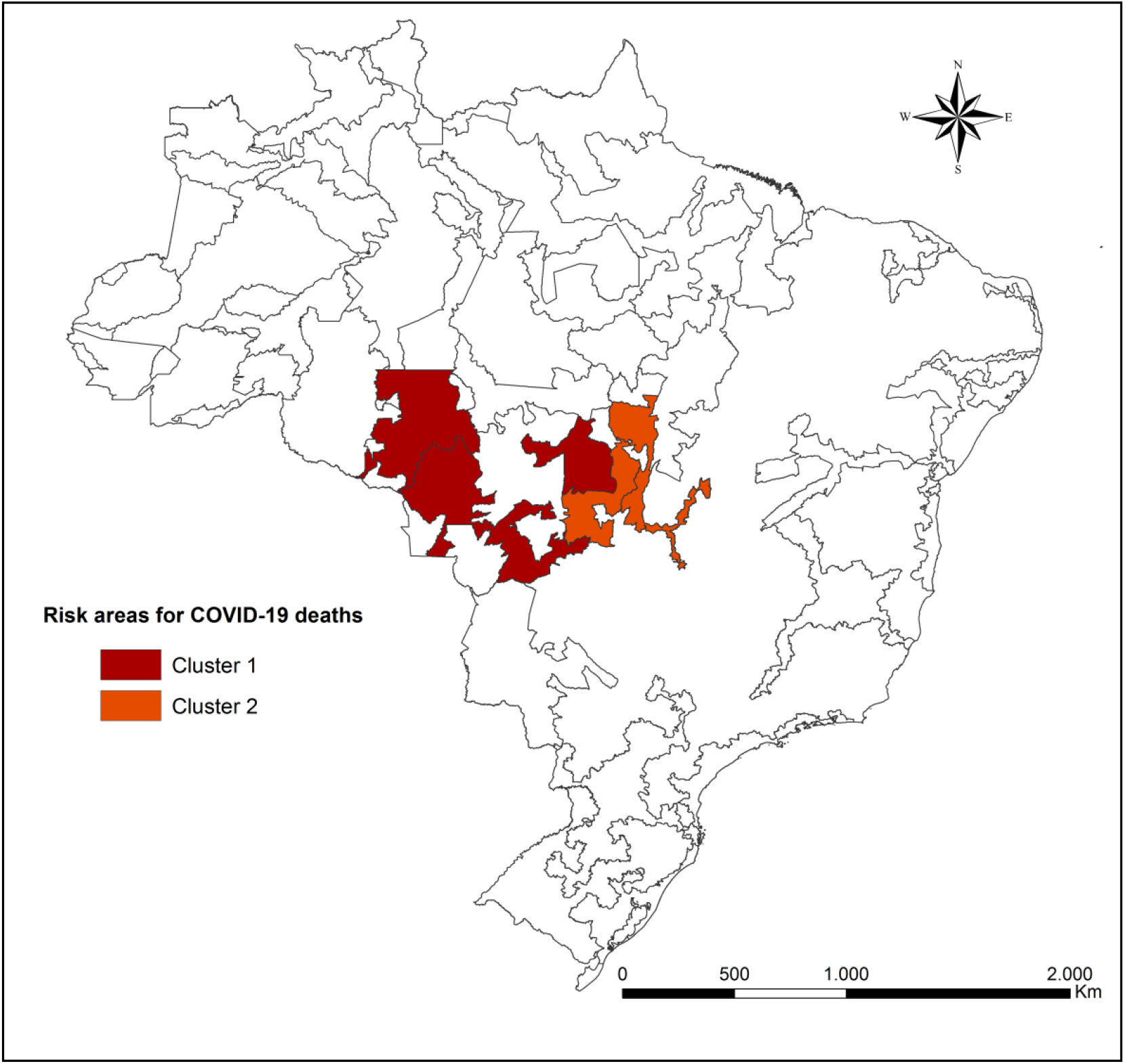
Spatial areas of risk of mortality due to COVID-19 in the indigenous population according to the special indigenous health district. Brazil, 2020.

## DISCUSSION

This study sought to verify the spatial distribution of COVID-19 in the indigenous population, and to identify areas of high risk for the occurrence of cases and deaths from the disease. The results demonstrated that COVID-19 is present in all SIHDs in the national territory and that there are high-risk spatial clusters for the investigated events, mainly in the Midwest and North of the country.

One of the limitations of this study is the use of secondary sources of information, which does not exclude the possibility of incomplete data. However, the information is obtained from each of the 34 SIHDs and, afterwards, is validated by the Department of Attention to Indigenous Health [10]. Another issue is that the data provided by the Ministry of Health are aggregated by SIHD, which includes several ethnic groups and villages within the same region. The provision of aggregated data in smaller geographical units, would allow a more detailed analysis, with the inclusion of variables related to sociocultural diversity and health determinants of these communities. The possibility of underreporting COVID-19 cases should also be considered.

Admittedly, the indigenous population has a high degree of vulnerability for both COVID-19 and other diseases, and our study showed that this population suffers strongly from the impacts of this worldwide pandemic, since it has indicators that far exceed those of the general population.

The results showed that the cases and deaths by COVID-19 peaked in July, and increased even in the face of the contingency plans launched by the government [23]. The incidence and mortality in the SIHDs for the period studied reached 18530.56 cases per 100000 people and 265.37 deaths per 100000 people, respectively, significantly exceeding the national rates registered in July, which were 1139.4 cases per 100000 people and 41.1 deaths per 100000 people [9,10].

Most of the deaths investigated occurred in males, following the worldwide trend, in which the majority of deaths occurred in the elderly male population[2]. Our findings also showed that incidence and mortality rates increase with the aging of the indigenous population, regardless of gender, which is in line with data from the general population of Brazil [9].

In order to better understand the impacts of the COVID-19 pandemic, it is necessary to consider some questions about vulnerabilities present in the Brazilian indigenous population. Firstly, these individuals already have a higher prevalence of infectious diseases, respiratory diseases, chronic diseases such as diabetes and hypertension, in addition to high rates of malnutrition and obesity [24]. Second, the social issue related to the source of income, with indigenous peoples being among the lowest income in the country [15]. Third, it should be noted that the indigenous population has many sociocultural specificities, which influence their relationship with the environment in which they live, their way of life and also the health-disease process. An example that can be mentioned is the high number of residents per household, sharing of personal utensils and collective ceremonies, such as weddings and funerals [25].

Fourthly, we highlight territorial vulnerability. A large part of the indigenous population suffers from the devastation of their lands due to the lack of demarcation, illegal invasion, mining and deforestation [26]. Finally, it is necessary to draw attention to the precarious health structures, lack of supplies, equipment, lack of training and high turnover of professionals, difficulty in removing patients who need assistance in medium and high complexity systems. In addition, health care is still focused on palliative and emergency practices, due to the lack of strengthening of primary care [27].

These issues became even more evident with the arrival of the COVID-19 pandemic and certainly contribute to the high rates of disease incidence and mortality among this population. The results of this study showed that there are clusters of risk for both, the occurrence of cases and deaths, which suggests that there are different degrees of vulnerability to COVID-19 among indigenous territories.

Azevedo et. al (2020) [28] constructed an index of demographic and infrastructural vulnerability of indigenous lands to COVID-19, which included variables related to the number of elderly people, number of residents per household, sanitary conditions of the household, proximity to places with availability of treatment beds, intensive and land tenure regularization of the indigenous land. The index varies between zero and one, the closer to one the greater the vulnerability. The SIHD with critical level of vulnerability (highest level) were: Alto Rio Negro, Yanomami, Xavante, Xingu, Kaiapó do Pará and Rio Tapajós. Coincidentally, these SIHD are part of the risk areas identified in our study. The SIHD in the Northeast and South of the country have a lower vulnerability index compared to the others, which corroborates the results of our study, that identified coldspots in these locations using the Gi^*^ analysis.

Two Brazilian regions stood out in the spatial analysis carried out in this study, the North and Midwest regions. The North region has a larger indigenous population compared to other regions. In this region, there is the state of Amazonas, which has the largest number of indigenous people in the country [15]. In addition, 10 of the 14 high-risk SIHDs for coronavirus infection were found in this study are located in the region.

Some regional specificities are challenging for the health care of the indigenous population of the North region, such as access to several villages, which is only possible by air or river, being affected by seasonal influences, especially during periods of drought in the rivers, increasing the travel time and late delivery of inputs. Work difficulties in these areas involve health risks, language barriers and poor housing conditions, favoring high professional turnover in the most remote regions [29].

The study by Silva et al (2016) [30] points out the main problems related to indigenous health in the Amazon region: scheduling appointments; long wait for hospital procedures (consultations, exams and surgeries); financial difficulty to buy medicines or to make private consultations and exams and the precarious physical structure of the services.

Another factor that may be favoring the transmission of COVID-19 in the North region is the number of residents per household, with households with up to 41 residents, approximately the double that is found in the South and Southeast regions. In addition, the indigenous population of the North region suffers from a high level of illiteracy, low income and poor sanitary conditions [15,24].

The Midwest region had a high incidence of COVID-19 and was the second region with the highest number of risk for the occurrence of the disease. The Midwest also draws attention on the national scene because it is an area with high mortality due to COVID-19. Our results showed that the two high-risk death clusters that were identified, belong to this region. Among the five SIHDs that were part of these clusters, the SIHD Xavante stands out, which is the most populous in the Midwest region and is located in the state of Mato Grosso. This SIHD differs from the others in that it is composed of only one ethnic group, totaling more than 21000 individuals [16].

Despite not belonging to the risk clusters of incidence of COVID-19, the SIHD Xavante was part of the risk cluster for deaths. In the month of July that was the peak of the pandemic, this SIHD reached the highest mortality rate and highest lethality among all Brazilian SIHDs, reaching 139.7 deaths per 100000 inhabitants with a lethality of 9.7% [10].

Although it is the most populous, the SIHD Xavante is the one with the most reduced structure in proportional terms. For each IBHU there are 670 indigenous people, while in the SIHD Xingu, the second most critical, that number drops to 348, almost half. Another worrying issue is that only 8.5% of Xavante villages have IBHU, that is, 91.5% of local communities do not have surveillance measures most of the time. [16,31].

It is not only the availability of the basic health care structure that positions the Xavante as one of the most vulnerable peoples to COVID-19 in the Midwest region. Aspects of their socio-cultural organization and, especially, of their epidemiological profile, marked by high rates of hypertension, diabetes mellitus and obesity, tend to increase the risk of developing severe forms of COVID-19 [32,33].

In addition, the form of organization of the Xavante villages, with very close houses and many residents, combined with the practice of collective events, such as the burial of their dead, may explain the high spread of COVID-19 in these territories [31].

In view of the above, the current vulnerability of these peoples is not only the result of an unforeseen pandemic, but part of a history of political and social exclusion that has historically violated their citizen rights and fundamental dimensions of their ways of life

High transmission of the virus combined with the low resolution of actions aimed at controlling the disease has culminated in a constant threat to life within indigenous communities. Although there is a national COVID-19 contingency plan for the indigenous population, the efforts made so far are reactive and late in nature. The control of the disease is difficult and becomes even more complex with the specificities of the indigenous population, requiring multisectoral actions. If control strategies are focused on areas of potential risk, it is likely that the impacts of COVID-19 will be mitigated and deaths will be avoided, especially in view of the current trend of reducing protection measures.

This study advances knowledge because it targets the indigenous population, considered one of the populations with the highest degree of vulnerability to COVID-19, as well as for using spatial analysis techniques as a tool to address this problem.

## CONCLUSION

This study allowed the identification of hot and cold areas for occurrence and deaths by COVID-19 in the Brazilian indigenous population. High-risk clusters were also identified for both events, which were mainly concentrated in the North and Midwest regions of the country. In this sense, we suggest that actions to combat COVID-19 in indigenous territories be strengthened and better targeted, considering mainly the risk areas identified in this study.

## Data Availability

The data that support the findings of this study are openly available in at https://coronavirus.saude.gov.br/boletins-epidemiologicos

## AUTHOR CONTRIBUTIONS

JDA and ASA: conceptualization; WPP, SMS, JEB: data curation; JDA, ASA, ARS: formal analysis; JDA and ARS: methodology; JDA and WPP: project administration; JDA, ASA, WPP, JEB and SMS: writing – original draft; WPP and ASA: Validation; JDA, ASA, WPP, JEB, SMS and ARS: writing – review & editing.

